# The Social Determinants of Depression among Adolescents in Low- and Middle-Income Countries: A Scoping Review

**DOI:** 10.1101/2025.04.07.25325376

**Authors:** Isayas Wubshet, Kibur Engdawork, Semere Gebremarim, Clementine Kanazayire, Pamela Abbott

**Affiliations:** Department of Sociology, Addis Ababa University, Addis Ababa, Ethiopia; Centre for Global Development, University of Aberdeen, UK; School of Medicine, Addis Ababa University, Addis Ababa, Ethiopia; College of Medicine and Health Sciences, University of Rwanda, Rwanda; School of Education, University of Aberdeen, UK

**Keywords:** Adolescent depression, social determinants, family, school, community, risk factors, protective factors, stressors, low- and middle-income countries (LMICs)

## Abstract

In Low and Middle Income Countries (LMICs), adolescent depression is one of the leading mental health problems with serious implications, posing a significant public health concern. A comprehensive understanding of adolescent depression and the development of effective intervention strategies require not only an examination of its biological and psychological determinants but also a critical analysis of the social factors influencing its prevalence and impact. This scoping review, guided by Arksey and O’Malley’s framework, examined the social factors influencing adolescent depression. A total of 48 primary studies were included, identified through a systematic search across five databases and manual exploration of relevant literature from various LMICs. This review identified structural factors including gender, socioeconomic status, ethnicity, and family structure alongside stressors in family, school, and neighborhood settings, such as conflict, harsh parenting, maltreatment, academic pressure, peer rejection, bullying, and neighborhood violence, as key contributors to adolescent depression. Conversely, social support from family, peers and teachers was found to be a protective factor against depression. However, these studies are primarily cross-sectional, with a predominant focus on Asia and limited attention to neighborhood-level factors, often concentrating on school-going adolescents. Furthermore, the studies are largely descriptive and atomized, inadequately exploring the domains and their interplay with social structures in shaping adolescent depression.

## 1. Introduction

Adolescence, spanning ages 10 to 19, is a critical developmental stage marked by significant biological, psychological, and social changes (Casey et al., 2010; Sreekanth et al., 2023; WHO, 2024a). These transitions expose adolescents to various social stressors (Petersen et al., 1991 as cited in Aseltine et al., 1994, p. 252), making this period particularly sensitive, vulnerable and challenging (Bodnar, 2022), and highly stressful (Sreekanth et al., 2023). Failure to effectively manage this increased sensitivity to stress can lead to heightened vulnerability to a range of internalized and externalized behavioral responses (Mukangabire et al., 2021; Sreekanth et al., 2023).

Adolescents constitute 16.7% of the global population (WHO, 2024b), with 90% of the world’s adolescent population residing in LMICs, as reported by the United Nations in 2019 (Sachin et al., 2023). Studies indicate that depression is the fourth most common mental health disorder among adolescents worldwide (Reangsing et al., 2024), affecting 14.3% of this population (WHO, 2024d).

Depressive symptoms, including sadness, loss of interest or pleasure, feelings of guilt or low self-worth, disturbances in sleep or appetite, fatigue, and poor concentration, commonly emerge during adolescence and it often persists into later life (Menaghan, 2010). Depression is frequently misidentified as an adult-specific condition (Bodnar, 2022), which can lead to adolescents’ symptoms going unnoticed by parents, teachers, and caregivers, delaying diagnosis and treatment (Sreekanth et al., 2023).

Depression extends beyond its immediate physical effects, such as immune system impairment (Arianti & Isnaeni, 2023), profoundly disrupting social relationships and academic performance. These disruptions can have lasting consequences, increasing the risk of social dysfunction, self-harm, and suicide (Curtis, 2015; Fergusson & Horwood, 2007; James et al., 2024; Menelik et al., 2017; Sreekanth et al., 2023; WHO, 2023).

Depression arises from a complex interplay of biological, psychological, and social factors. A comprehensive understanding and effective management of this condition require an enhanced focus on social determinants alongside the well-established biological and psychological foundations (NRCIM, 2009; Remes et al., 2021; WHO, 2023, 2024c). The WHO (2024e) defines social determinants as “the conditions in which people are born, grow, work, live, and age, and the wider set of forces and systems shaping the conditions of daily life.”

Unlike biomedical and psychological approaches, which primarily focus on physiological and internal factors, sociological perspectives underscore the significance of social integration, social stratification, inequality, and cultural values as critical determinants that can contribute to the risk of adolescent depression (Mirowsky & Ross, 2003). In this regards, the stress process theory provides a comprehensive framework for understanding how social conditions contribute to individual stress (Pearlin, 1982, 1989; Thoits, 2010).

Social stress theory posits that external social factors influence individuals through their immediate social environments, shaping their exposure to stressors. The accumulation of life events and chronic social stressors can elevate stress levels, ultimately increasing the risk of depression. Social support plays a crucial role in this process, serving as a protective factor that mitigates the negative effects of stressors and reduces vulnerability to depressive symptoms. Conversely, insufficient social support from one’s immediate social network can exacerbate stress, increasing the likelihood of depression (Aneshensel & Mitchell, 2014; Pearlin & Bierman, 2013; Pearlin et al., 1981; Wheaton, 2010).

Adolescents are situated within various social environments that play a crucial role in shaping their development. As Bronfenbrenner suggests, the most immediate social contexts—family, school, and neighborhood—are integral to their overall well-being and developmental outcomes (Bronfrenbrenner, 1979). The family plays a crucial role in the transition from childhood to adulthood, providing essential emotional and social support that significantly influences adolescents’ development (Bronfenbrenner, 1986; Shao et al., 2020; Wight et al., 2006). Schools are key socializing institutions where adolescents form peer relationships and build connections with teachers and staff, shaping their daily lives through institutional structures, norms, and values. Neighborhoods play a key role in adolescent development by facilitating peer interactions and providing access to social engagement and community resources beyond the family and school (Boardman & SaintOnge, 2005). Additionally, the growing prevalence of social media has further reshaped adolescent socialization, with many adolescents using smartphones to cultivate friendships and navigate the virtual world (Ekbäck et al., 2024; Radovic et al., 2017).

Social structural influences introduce various social stressors, significantly impacting adolescents’ well-being. Adolescents from low socioeconomic status, single-parent, or divorced families are more vulnerable to depression (Laukkanen et al., 2016; Waldfogel et al., 2010; Zeiders et al., 2011). Maltreatment, family conflict, and strained parent-adolescent relationships are significant family-related stressors that contribute to the development of depressive symptoms (Cheng et al., 2015; Patricia et al., 2021; Xu et al., 2019). School-related stressors, including bullying and academic pressures, also increase the risk of depression (Frojd et al., 2008). Additionally, adolescents in neighborhoods with crime and violence face greater risks (Cho, 2022) and social media use has also been linked to higher depression levels in adolescents (Ekbäck et al., 2024; Radovic et al., 2017).

Adolescents rose in stable family environments with positive relationships and strong parental support are less likely to experience depression (Mohanraj et al., 2010; Nduwimana, 2017; Pekcanlar et al., 2006; Wang, Sun, et al., 2023; Wang, Ting Zhou, et al., 2023; Yan et al., 2014). Similarly, positive relationships with teachers, peer acceptance, strong academic performance, and a supportive school environment act as protective factors, promoting adolescent well-being (Hunduma et al., 2022; Mohanraj et al., 2010; Ong et al., 2015). Additionally, a supportive neighborhood and strong community social networks can serve as protective factors against depression (Chung et al., 2020; Mori, 2022).

Despite numerous studies, the social factors shaping adolescent depression remain inadequately understood. Grounded in Pearlin’s stress process theory, this review synthesizes existing research, particularly from low- and middle-income countries (LMICs), to examine how external structural factors and proximate social contexts shape adolescents’ depressive experiences. By identifying gaps in the literature, this analysis aims to inform future research and public health strategies.

## 2. Method

The purpose of this study was to systematically map research investigating the social determinants of adolescent depression within the context of LMICs. A detailed protocol for this scoping review was pre-published on MedRxiv (Isayas et al., 2024). The review process was conducted using Covidence, and a summary of the key modifications made to the protocol is provided below.

### 2.1. Research Question

The primary objective of this review was to examine the social determinants of depression among adolescents in LMICs. Specifically, it aimed to explore how social factors, mediated through social contexts such as family, school, and neighborhood environments, contribute to stress and subsequently lead to depression, with social support acting as an intermediary. Additionally, the review sought to identify gaps in the existing literature to inform future research in this area.

### 2.2. Eligibility Criteria

This review included empirical studies published in English between January 2010 and December 2023 that focused on depression among adolescents residing in LMICs. Studies using any research methodology were considered, provided they included a general sample (excluding adolescents with diagnosed biological or psychological conditions) and examined risk/protective social factors influencing adolescent depression.

### 2.3. Data Source and Search Strategy

The primary databases searched included Google Scholar, PsychINFO (via Ovid), Web of Science, PubMed, Sociological Abstracts (via ProQuest), and Embase (via Ovid). A set of search terms was employed to encompass all potential terminology related to adolescent depression, with the full list detailed in the protocol. Search results were organized using EndNote reference management software.

### 2.4. Screening and Data Extraction

An initial search using terms approved by PA, IW, and SG retrieved 48,468 records. After removing 42,045 duplicates with Covidence, 6,417 records remained for title and abstract screening. IW and SG excluded 5,964 irrelevant records, leaving 453 for full-text review. Additionally, 16 studies were identified through manual searches using backward citation and a Google search of the review’s title, and were directly included in the full-text assessment.

Before the full-text review, IW and SG independently screened a sample of retrieved records to ensure consistent application of eligibility criteria, achieving a 94% inter-rater agreement, which exceeded the 80% threshold (McHugh, 2012). Discrepancies were resolved through discussion. Of the 453 potentially relevant articles, 48 were included in the review. All the included studies had been peer reviewed.

Key data, including study design, sampling method, data collection, and findings, were extracted by IW and verified by SG, KE, and CK. The article selection process, following PRISMA guidelines, is presented in Figure 2.

**Fig 1:**
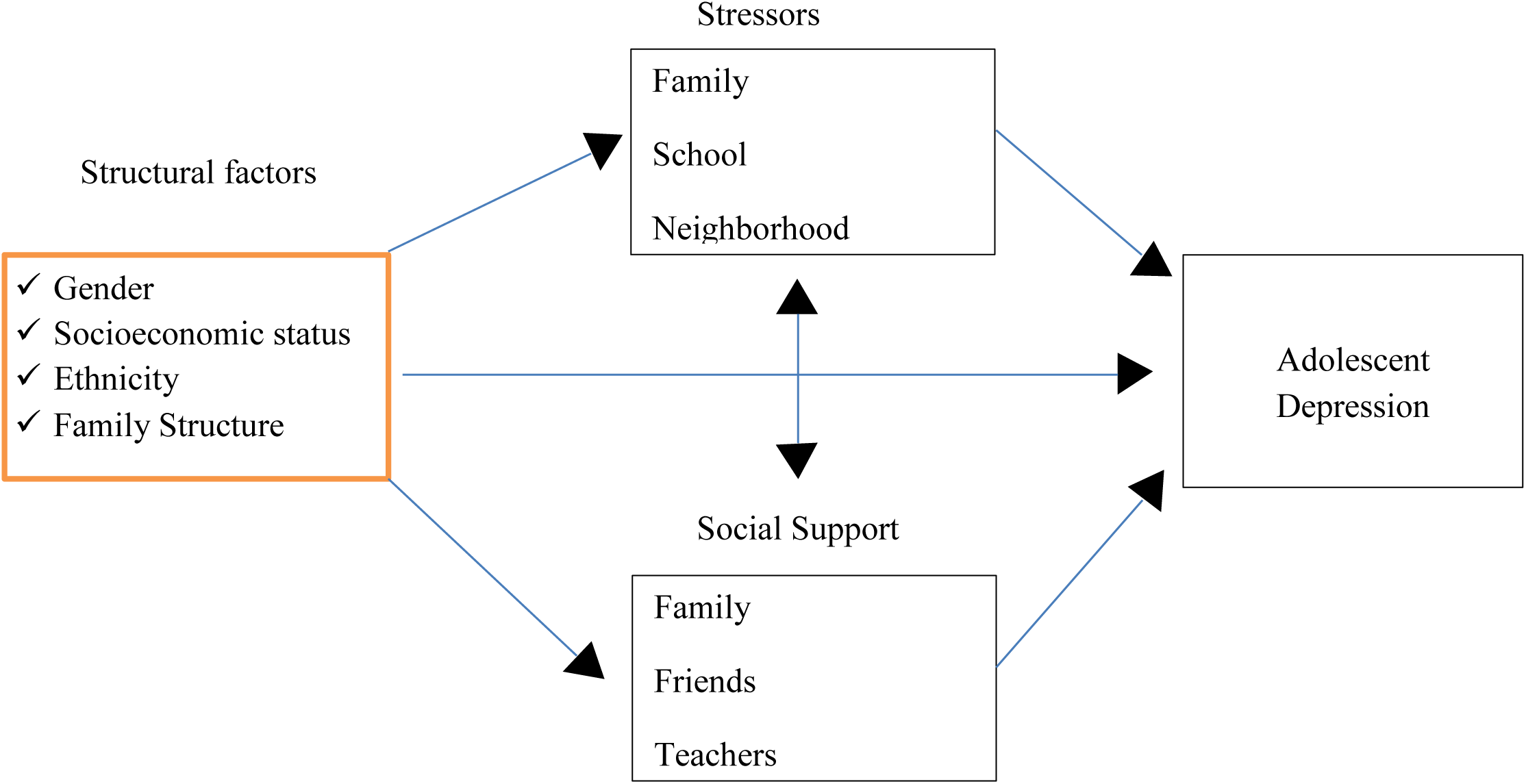
Conceptual framework of the study (based on Pearlin’s stress process theory).

**Fig 2:**
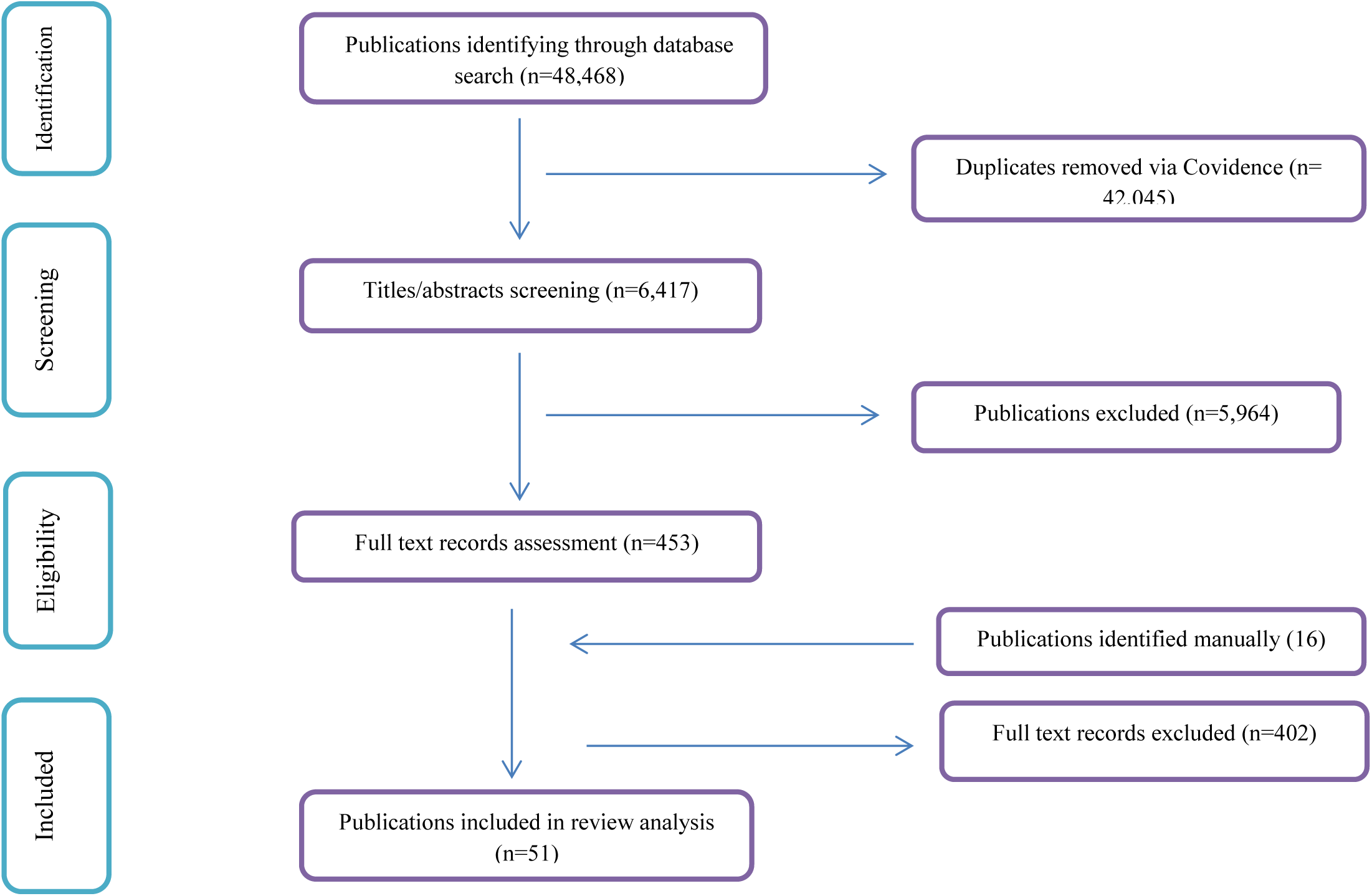
The PRISMA flow diagram.

The selected records were uploaded into ATLASt.ti Version 9, where IW and SG independently coded a randomly selected 10% sample. Following feedback from PA and KE, IW and SG proceeded to independently code the remaining records. To ensure consistency and reliability, regular discussions were held to address any coding discrepancies and review the content of the codebook. These meetings facilitated the alignment of coding approaches and the comparison of results.

### 2.5. Changes to the Pre-Published Protocol

There were some modifications made to this review compared to the pre-published protocol. First, the original protocol specified an age range of 10–18 years for included adolescents. However, during the review process, it became apparent that the definition of adolescence varied across studies. Some studies defined participants by mean ages (e.g. Alex et al., 2023; Ng & Wan, 2017), others included individuals older than 18 years (e.g. Liu, 2023; Wahid et al., 2022), and some did not specify participants’ ages (e.g. Jayanthi, 2015). As a result, rather than strictly adhering to the proposed age range, we included studies that focused broadly on adolescents while excluding those involving children under 10. Second, while the original protocol limited the scope to studies conducted in urban settings, the review was expanded to include studies from both urban and rural areas, provided the research did not focus exclusively on rural settings and included an urban component. Third, although the protocol initially planned to include gray literature, the review was restricted to academic literature. This decision was due to the broad scope of the review, the challenges of incorporating country-specific gray literature, and concerns over potential biases. Finally, while the protocol emphasized searching specific databases, an initial assessment of titles and abstracts revealed that many studies originated from China and predominantly focused on the family domain. To address this imbalance and achieve a more comprehensive review, manual searches were conducted with other team members to identify additional studies, particularly those examining school and neighborhood domains, for full-text review.

## 3. Results

### Characteristics of Included Studies

Out of the 48 studies included in this review, more than half—25 studies (52.1%)—were conducted in China. Five studies (10.4%) were from India, three from Malaysia, and two each from Ethiopia, Kenya, Nepal, and Pakistan. The remaining seven studies were from seven different countries: Albania, Brazil, Cambodia, Indonesia, Jamaica, Macedonia, and South Africa. When considering the publication dates, 33 studies were published in 2020 or later, while the remaining 9 studies were published between 2015 and 2019 and the remaining 6 studies were published in between 2010-2014.

Of the 48 studies included, 40 were cross-sectional, while eight were longitudinal or panel studies. Only one employed a qualitative research approach (Wahid et al., 2022). Of the remaining 47 quantitative studies, 46 relied on probability sampling techniques to select respondents, with only two (Shi et al., 2023; Xia, 2022) using non-probability sampling methods.

Although all the studies focused on adolescents, three also included other groups: parents (Emery et al., 2022; Wahid et al., 2022), siblings (Ng, 2017), as well as teachers and professionals (Wahid et al., 2022). Furthermore, while all studies examined school-going adolescents, two studies (Miloseva et al., 2017; Wahid et al., 2022) specifically involved adolescents with a history of depression who were undergoing treatment in psychiatric clinics. In total, this review analyzed data from 108,409 participants, with sample sizes ranging from 72 to 14,500, of whom 99.9% (108,349) were adolescents.

Twelve studies used the Center for Epidemiologic Studies Depression (CES-D) Scale to screen for adolescent depression (Emery et al., 2022; Guo et al., 2014; Lakhdir et al., 2021; Miloseva et al., 2017; Pan et al., 2022; Peng et al., 2021; Shen et al., 2022; Singh et al., 2023; Wang, Sun, et al., 2023; Xu et al., 2019; J. Zhang et al., 2022; Zou et al., 2020). Additionally, in nine studies (Bach & Louw, 2010; Dervishi et al., 2019; Dhamayanti, 2020; Khasakhala et al., 2012; Liu, 2020; Qin, 2021; Song et al., 2022; Vasconcelos, 2020; Xia, 2022), the Children’s Depression Inventory (CDI) was employed. The Beck Depression Inventory (BDI) (Jayanthi et al., 2016; Jayanthi, Thirunavukarasu, & Rajkumar, 2015; Kaur & Sharma, 2014; Liu et al., 2022; Mohanraj et al., 2010; Ng, 2017) and the Patient Health Questionnaire-9 (PHQ-9) (Aklile et al., 2023; Ibrahim et al., 2017; Li et al., 2022; Mekonen et al., 2020; Naveed et al., 2019; Pan et al., 2022) were each used in six studies to screen for adolescent depression. These are followed by the Depression Self-Rating Scale for Children (DSRSC) (Liu, 2023; Shi et al., 2023) and the Feeling of Four-Item Scales for the Past Seven Days (Wang et al., 2022; Zhang et al., 2022), which were each used in two studies. The remaining 11 studies employed other psychometric tools to screen for depression.

### Age

Studies in this review examined the relationship between age and adolescent depression using two approaches. Five studies directly measured the association between age and depressive symptoms, reporting a positive correlation that indicated older adolescents experienced higher levels of depression (Dervishi et al., 2019; Emery et al., 2022; Khasakhala et al., 2012; Naveed et al., 2019; Sun et al., 2021). On the other, there are studies which used grade level as a proxy for age. Consistent with the first approach, these studies found that high school students reported higher levels of depression compared to middle school students (Guo et al., 2014; Xu et al., 2019). Similarly, middle school students exhibited higher levels of depression compared to primary school students (Wang, Ting Zhou, et al., 2023). Overall, the findings suggest a positive association between increasing age and depressive symptoms during adolescence (He et al., 2019; Ibrahim et al., 2017; Li et al., 2022; Ma et al., 2020; Mekonen et al., 2020; Miloseva et al., 2017; Paulo et al., 2022; Xu et al., 2019).

### Gender

Almost half of the studies in this review examined the potential association between gender and depressive symptoms. Studies indicated that girls tend to report higher levels of depression compared to boys (Aklile et al., 2023; Bach & Louw, 2010; Chung et al., 2020; He et al., 2019; Jayanthi, Thirunavukarasu, & Rajkumar, 2015; Khasakhala et al., 2012; Lakhdir et al., 2021; Li et al., 2022; Ma et al., 2020; Mekonen et al., 2020; Miloseva et al., 2017; Pan et al., 2022; Paulo et al., 2022; Peng et al., 2021; Singh et al., 2023; Song et al., 2022; Sun et al., 2021; Wang, Sun, et al., 2023; Wang, Ting Zhou, et al., 2023; Xu et al., 2019; Yi et al., 2013; Zhang et al., 2022; J. Zhang et al., 2022).

Girls from low socioeconomic backgrounds (Miloseva et al., 2017), with low parental education (Qin, 2021; Sun et al., 2021; Yi et al., 2013), or from single-parent households (Sun et al., 2021) are more likely to experience depression than boys. Family-related stressors, such as poor paternal and maternal relationships (He et al., 2019; Peng et al., 2021; J. Zhang et al., 2022), family conflict or violence (Li et al., 2022; Miloseva et al., 2017; Paulo et al., 2022), high maternal control(Xu et al., 2019), and adverse childhood experiences like abuse, maltreatment, and negative life events (Lakhdir et al., 2021; Paulo et al., 2022; Zhang et al., 2022), increase the risk of depression among girls. Similarly, girls experience higher levels of school-related stressors, including bullying (Metasebia, 2020; J. Zhang et al., 2022), academic pressure (Ma et al., 2020; Pan et al., 2022; Sun et al., 2021), weak school connectedness (He et al., 2019), and low peer support (Miloseva et al., 2017). Additionally, neighborhood-related stressors, such as witnessing violence (Li et al., 2022), further elevate depression rates among adolescent girls.

### Socioeconomic Status

Socioeconomic status (SES) as a potential determinant of adolescent depressive symptoms was assessed using both objective and subjective measures. Objective measures included family income (Khasakhala et al., 2012; Xu et al., 2019; Yi et al., 2013), home ownership (Yi et al., 2013), material possessions (Zou et al., 2020), and parental education and employment status (Lakhdir et al., 2021; Xu et al., 2019). Additionally, two cross-sectional and two longitudinal studies conducted in China utilized subjective measures, which relied on self-reported ratings or rankings of SES (Guo et al., 2014; Pan et al., 2022; Qin, 2021; Zhang et al., 2022). With the exception of study conducted in China involving middle school adolescents (Qin, 2021) which showed no association, all other studies found a negative association between socioeconomic status and adolescent depression (Guo et al., 2014; Khasakhala et al., 2012; Lakhdir et al., 2021; Pan et al., 2022; Xu et al., 2019; Yi et al., 2013; Zhang et al., 2022; Zou et al., 2020). This is because families with higher socioeconomic status (SES) tend to foster stronger parent-child relationships (Wang, Ting Zhou, et al., 2023; Zhang et al., 2022), experience lower school-related stress (Wang, Ting Zhou, et al., 2023), receive greater social support (Zou et al., 2020), and benefit from improved maternal care (Xu et al., 2019). Contrary to this, an Indian study conducted on girls from two private and two public schools found that adolescents with educated parents were nearly three times more likely to experience depression than those whose parents had no formal education (Alex et al., 2023).

### Ethnicity

Two studies examined ethnicity as a social factor influencing depression. Research conducted in China found a significant association between ethnicity and depressive symptoms (Liu, 2020). Similarly, a study in South Africa revealed that Venda adolescents experienced higher rates of bullying, violence, and mistreatment compared to their Northern Sotho peers. Despite facing greater victimization, Venda adolescents exhibited lower rates of depression than Northern Sotho boys. Regardless of ethnicity or gender, individuals exposed to violence, including bullying and physical attacks, were more likely to develop depressive symptoms (Bach & Louw, 2010).

### Family Structure

This review highlights studies suggesting that family type and size significantly influence the risk of adolescent depression. Cross-sectional studies conducted in three Asian countries (China, India, and Cambodia) have shown that adolescents living in nuclear families are more likely to experience depression compared to those living in extended family households (Alex et al., 2023; Sun et al., 2021; Yi et al., 2013). Similarly, a study conducted in Ethiopia found that adolescents from smaller families were three times more likely to develop depression compared to those from larger families (Aklile et al., 2023). Adolescents from larger or extended families often benefit from robust social networks and emotional support, which serve as protective factors against depression by providing coping mechanisms to manage stressors (Aklile et al., 2023; Alex et al., 2023). While adolescents with siblings have been found to have higher rates of depression compared to only children, this association is not robust when positive parent-adolescent relationships are present (Song et al., 2022).

### Family Stressors

Adolescents exposed to life events such as parental illness, alcohol use, death, divorce, and related hardships, as well as stressful daily events, face a higher risk of developing depressive symptoms compared to their peers (Alex et al., 2023; Gao, 2022; Jayanthi et al., 2016; Lakhdir et al., 2021; Liu, 2020; Mekonen et al., 2020; Mohanraj et al., 2010; Pan et al., 2022; Paulo et al., 2022; Xia, 2022).

Except for a Chinese longitudinal study involving primary school adolescents (Wang et al., 2022), which found no association with parent-adolescent communication, studies consistently showed that positive family relationships—including closeness, a positive family atmosphere, positive parental behavior, and social support—act as protective factors, improving coping mechanisms and reducing the risk of depressive symptoms (Huang, 2022; Li et al., 2022; Liu et al., 2022; Ma et al., 2020; Mohanraj et al., 2010; Ng, 2017; Shi et al., 2023; Wang, Sun, et al., 2023; Wang, Ting Zhou, et al., 2023; Xu et al., 2019; Yi et al., 2013; Zhang et al., 2022; J. Zhang et al., 2022; Zou et al., 2020).

Conversely, high levels of family conflict emerged as one of the most significant risk factors for adolescent depression. Studies consistently demonstrated that conflict between adolescents and their parents (He et al., 2019; Hua & Liu, 2021; Wahid et al., 2022; Wang, Sun, et al., 2023; Wang et al., 2022), as well as conflict among other family members (Alex et al., 2023; Li et al., 2022; Wang, Ting Zhou, et al., 2023; Yi et al., 2013), and limited family social support (Gao, 2022; Yi et al., 2013) significantly increased the risk of depressive symptoms. Notably, girls appeared more susceptible to the negative impacts of parental quarrels and emotional punishment, particularly from mothers (He et al., 2019) and fathers (Peng et al., 2021).

Studies from three Asian countries—China (He et al., 2019; Hua & Liu, 2021; Qin, 2021; Xu et al., 2019), Cambodia (Yi et al., 2013), and Malaysia (Singh et al., 2023)—as well as a study conducted in Ethiopia (Aklile et al., 2023), found that adolescents with greater parental autonomy experienced lower levels of depressive symptoms compared to those with psychologically controlling parents (Qin, 2021). Negative parenting practices, including physical and emotional punishment by parents (Yi et al., 2013), maternal rejection or under protective behavior (Khasakhala et al., 2012), high maternal control (Xu et al., 2019), lack of parental supervision (Aklile et al., 2023; Singh et al., 2023), and parenting through lying (Hua & Liu, 2021), were shown to significantly increase the risk of depressive symptoms.

High exposure to adverse childhood experiences (ACEs) has been linked to increasing adolescents’ susceptibility to depressive symptoms (Paulo et al., 2022). For instance, studies in China (Song et al., 2022), Kenya (Paulo et al., 2022), South Africa (Bach & Louw, 2010) and Ethiopia (Aklile et al., 2023) have found that adolescents with a history of abuse were more than twice as likely to experience depression. Similarly, a qualitative study from Nepal supported this finding, indicating that adolescents with a history of neglect and sexual abuse exhibited depressive symptoms (Emery et al., 2022). An Indonesian study reported an even higher risk, with adolescents who experienced abuse being more than 6.5 times more likely to be depressed (Dhamayanti, 2020). Longitudinal studies from Pakistan (Lakhdir et al., 2021) and China (J. Zhang et al., 2022) revealed that maltreated or abused girls were at a significantly higher risk of depression compared to boys, with the risk more than doubling (J. Zhang et al., 2022). Psychological child abuse was found to be a more significant predictor of early-onset depression compared to physical or sexual abuse (Vasconcelos, 2020; J. Zhang et al., 2022).

### School Stressors

Studies show a positive link between academic pressure from teachers and families and the development of depressive symptoms in adolescents (Alex et al., 2023; Kaur & Sharma, 2014; Liu, 2020; Miloseva et al., 2017; Pan et al., 2022). Poor academic performance increase the risk of depressive symptoms, particularly in higher grade levels (Yi et al., 2013), while high-achieving students report fewer such symptoms (Qin, 2021).

Academic stress contributing factors include excessive test schedules (Alex et al., 2023), attending extracurricular tutoring sessions after school hours (Pan et al., 2022), and experiencing academic frustration (Kaur & Sharma, 2014) significantly increase the risk of depressive symptoms. Comparing to their peers with lower stress levels adolescents experiencing high levels of academic stress were two to four times more likely to exhibit depressive symptoms (Jayanthi, Thirunavukarasu, & Rajkumar, 2015).

Lack of peer acceptance, the absence of positive relationships, and a lack of supportive peers at schools significantly increase the risk of depression in adolescents (Alex et al., 2023; He et al., 2019; Wahid et al., 2022; Wang, Ting Zhou, et al., 2023). Adolescents who have been bullied are 3.7 times more likely to experience depressive symptoms compared to their peers (Singh et al., 2023). Prolonged exposure to bullying victimization (Naveed et al., 2019; Shen et al., 2022), especially for those who frequently engage in bullying behaviors (Dervishi et al., 2019; Ibrahim et al., 2017; Liu, 2023; Naveed et al., 2019; Singh et al., 2023; J. Zhang et al., 2022), significantly increases the risk of developing depressive symptoms. Boys, in particular, are at an even greater risk due to higher rates of victimization (Liu, 2023). Furthermore, research suggests that both victims and perpetrators of bullying are at increased risk of developing depression (Dervishi et al., 2019; Ibrahim et al., 2017).

Adolescents who face conflicts with their teachers or experience mistreatment and punishment are at a higher risk of developing depressive symptoms (He et al., 2019). Similarly, insufficient teacher support can also contribute to higher rates of depression among adolescents (Miloseva et al., 2017).

A longitudinal study in India found that students in private schools with high academic expectations and intense competition were more likely to exhibit depressive symptoms, whereas public school students faced less academic stress and lower risks of depression (Alex et al., 2023). However, other studies conducted in India (Jayanthi, Thirunavukarasu, & Rajkumar, 2015), and China (Aklile et al., 2023; Guo et al., 2014) challenge this finding, indicating that attending a government school may increase the risk of depression. While parents often see boarding schools as beneficial for discipline, the separation from family has been linked to a higher risk of depression among students (Khasakhala et al., 2012).

### Social Media Stressors

Studies indicated that adolescents who engage extensively in online gaming and watching TV (Pan et al., 2022) are more likely to develop depressive symptoms due to prolonged screen time and reduced social interaction. Similarly, excessive use of social media (Song et al., 2022) and smartphones (Alex et al., 2023) has been associated with an increased risk of depression.

### Neighborhood Stressors

While research examining the impact of neighborhood context on adolescent depression is limited, this review includes two relevant studies. A cross-sectional study conducted in Jamaica investigated the association between adolescents’ perceptions of their communities in terms of violence and their likelihood of experiencing depression. The results showed that positive neighborhood interactions were associated with a lower likelihood of exhibiting depressive symptoms. Notably, this study found that public school students, who often come from low socioeconomic status neighborhoods with higher levels of crime, violence, and other forms of adversity, were more vulnerable to depression compared to their peers attending private schools (Chung et al., 2020). Similarly, another study conducted in Cambodia found that being a victim or witness of community violence was positively associated with adolescent depression (Yi et al., 2013).

### Social Supports

Adolescents who receive strong social support from family and friends have a significantly lower risk of developing depression (Gao, 2022; Liu, 2020; Song et al., 2022; Xia, 2022). Compared to boys and children without siblings, girls and those with siblings who maintain strong relationships with their parents exhibit fewer depressive symptoms (Song et al., 2022). Similarly, high levels of peer acceptance (Mohanraj et al., 2010) and positive peer relationships (Wahid et al., 2022) are associated with reduced depressive symptoms. Furthermore, adolescents who have positive relationships with their teachers and receive sufficient support are less likely to experience depression than their peers (Gao, 2022; He et al., 2019; Liu et al., 2022; Wang, Ting Zhou, et al., 2023; Xu et al., 2019; Zou et al., 2020).

## 4. Discussion

The majority of studies included in this review were conducted in Asia (39), with a significant portion originating from China (25). In comparison, a smaller number of studies were carried out in Africa (5) and Europe (2), with only one study each from North America and South America. While Asia dominated the research landscape, the heavy concentration of studies from China limits the generalizability of the findings to the entire continent. Similarly, the representation from other continents remains minimal.

Most studies were conducted in upper-middle-income countries, including Albania, Brazil, China, Jamaica, Macedonia, Malaysia, and South Africa, followed by lower-middle-income countries such as Cambodia, India, Indonesia, Kenya, Nepal, and Pakistan. In contrast, research on adolescent depression in low-income countries was limited, with studies primarily focused on Ethiopia and Nepal, highlighting a significant gap in the literature on social determinants of adolescent depression in these settings.

The majority of studies were cross-sectional quantitative studies, with only eight longitudinal studies. Of these longitudinal studies, seven were conducted in China and one in Pakistan. Only one study, conducted in Nepal, employed a qualitative research methodology. The limited number of longitudinal and qualitative studies, particularly outside of China, represents a gap in the existing literature.

Studies have identified a positive association between increasing age and depressive symptoms in both high-income countries, such as the United States (Tepper et al., 2008) and South Korea (Zhang J et al., 2023), and low- and middle-income countries (Mekdes et al., 2018; Sandal et al., 2017; Shimelis et al., 2021). Similarly, extensive research (Aklile et al., 2023; Lin et al., 2008; Partap et al., 2023; Sandal et al., 2017; Shimelis et al., 2021; Wirback et al., 2014; Yan et al., 2014) and systematic reviews (Bazargan et al., 2023) consistently demonstrate that girls are more vulnerable to depression than boys, often attributed to their increased exposure to stressful life events and victimization. The findings of this review corroborate these results, revealing a strong association between increasing age and higher levels of depression in more than half of the studies, including those conducted across countries with varying income levels. Moreover, girls were consistently reported to be at a higher risk of experiencing depression compared to boys. Notably, longitudinal studies provided stronger evidence supporting the relationship between gender and adolescent depression compared to studies examining age as a factor.

The majority of studies indicate that low SES is a significant risk factor for adolescent depression. This association is attributed to various interconnected factors, including increased likelihood of resource deprivation (Atilola, 2014), disruptions in effective parenting (Menaghan, 2010), exposure to abuse and traumatic events (Aklile et al., 2023; Fletcher, 2009; Yi et al., 2013), financial stress leading to family conflict (Aklile et al., 2023; Ong et al., 2015; Sternberg et al., 2006), residence in disadvantaged neighborhoods (Latkin & Curry, 2003), and exposure to harsh parenting practices (Conger et al., 2010).

The reviewed studies suggest that adolescents from nuclear or smaller families are at a higher risk of experiencing depression. Research conducted in high-income countries (Kehusmaa et al., 2022; Laukkanen et al., 2016) has demonstrated that, beyond nuclear and small families, diverse family structures—including stepfamilies, blended families, and extended families— significantly influence adolescent depression. Furthermore, transitions within family structures, such as divorce or remarriage, have been shown to have substantial impacts on adolescent mental health. However, this review provides limited evidence on the effects of these diverse family structures and the impact of family structure transitions in LMICs.

The majority of studies in this review examined the relationship between depression and social stressors that adolescents experience within their family, school, neighborhood, and social media. Consistent with previous research (Ekinci, 2024; Grover et al., 2019; Kempfer et al., 2017; Kinyanda et al., 2013; Laukkanen et al., 2016; Lindert et al., 2014; Liranso & Singhe, 2018; Littler, 2016; Liu et al., 2022; Mackenbach et al., 2014; Mekdes et al., 2018; Musisi et al., 2007; Ong et al., 2015; Ozturk et al., 2021; Radell et al., 2021; Rizvi & Najam, 2014; Waldinger et al., 2007; Wirback et al., 2014; Ying et al., 2018) the findings of this review indicate that Adverse Childhood Experiences (ACEs), such as neglect, family conflict, and exposure to stressful life events, are consistently linked to an increased risk of depression. However, certain non-traditional family risk factors—such as parental migration, caregiving by non-parental guardians, intergenerational trauma, and parental mental health (Marshall, 2014; Sim, 2022; Wang, Ting Zhou, et al., 2023) remain underexplored in existing research.

Research has established a strong correlation between academic-related stress, academic pressure, and depressive symptoms among adolescents (Frojd et al., 2008; Grover et al., 2019; Jepper et al., 2008; Liu et al., 2024). Consistent with these findings, this review indicates that increased academic demands and heightened expectations from parents and teachers, particularly as adolescents advance to higher grade levels, are associated with a greater risk of depression. Furthermore, peer rejection, bullying, and negative interactions with teachers have been linked to a higher prevalence of depressive symptoms. Adolescents from low socioeconomic backgrounds and those exposed to highly stressful academic environments appear to be particularly vulnerable.

Systematic reviews highlight that neighborhood conditions, including safety, discrimination, and community violence, are significant predictors of depressive symptoms (Miliauskas et al., 2022; Stirling et al., 2015). Findings from two studies included in this review suggest that adolescents living in neighborhoods characterized by negative social interactions, low socioeconomic status, and high levels of violence, whether experienced directly or witnessed, are at an increased risk of developing depressive symptoms. However, the limited number of studies in this area highlights the need for further research to examine a broader range of neighborhood factors influencing adolescent mental health.

Conversely, positive parent-adolescent relationships and robust social support systems have been identified as protective factors, significantly reducing the likelihood of depression. These findings are supported by extensive research conducted in both high-income and low- and middle-income countries (Ekinci, 2024; Grover et al., 2019; Kempfer et al., 2017; Kinyanda et al., 2013; Laukkanen et al., 2016; Lindert et al., 2014; Liranso & Singhe, 2018; Littler, 2016; Liu et al., 2022; Mackenbach et al., 2014; Mekdes et al., 2018; Musisi et al., 2007; Ong et al., 2015; Ozturk et al., 2021; Radell et al., 2021; Rizvi & Najam, 2014; Waldinger et al., 2007; Wirback et al., 2014; Ying et al., 2018).

A comprehensive understanding of the social determinants of adolescent depression requires examining the complex interplay of factors within each domain (Carlén, 2022; Mijs & Nieuwenhuis, 2022). However, most studies have investigated these factors in isolation, with few exploring the combined effects of multiple social factors in family and school domains. This limitation restricts the understanding of the intricate interactions between various social determinants and adolescent depression. Additionally, the exclusive focus on school-going adolescents overlooks the experiences of out-of-school adolescents, who may be at a heightened risk of depression, particularly in LMICs.

## 5. Conclusion

This review aimed to examine the association between social factors and adolescent depression, with a particular focus on LMICs. It synthesized findings from 51 studies identified through both database and manual searches. Of these, 76% investigated family-related factors, 48.1% examined school-related factors, and only two studies explored neighborhood-related factors, highlighting it as the least studied area in the literature.

The review reveals significant regional and methodological gaps, with the majority of studies conducted in Asia, particularly China, and minimal representation from Africa, Europe, and South America. Most studies are cross-sectional, with few employing longitudinal designs, and only one qualitative study, highlighting the need for more diverse and robust study methodologies. Additionally, the exclusive focus on school-going adolescents leaves the experiences of out-of-school youth largely unexplored.

The studies in this review primarily focused on how social stressors from external environments contribute to adolescent depression. As a result, they overlooked sociological explanation on social determinants, particularly structural factors such as socioeconomic inequalities and cultural values, which play a fundamental role in shaping these social stressors and the development of depressive symptoms. Therefore, this simple description and atomization have limited the depth of analysis, potentially contributing to an incomplete understanding of the social determinants of adolescent depression.

This review emphasizes the need for research that explores how social factors contribute to depressive symptoms among adolescents by influencing their social circles. Rather than atomizing social factors and stressors, it is important to examine the interplay of multiple factors for a better understanding of the social determinants of adolescent depression in LMICs. Addressing these gaps is essential for developing targeted, context-specific interventions that effectively address the unique challenges faced by adolescents in diverse settings.

## 6. Limitation and Future Research

This study primarily emphasized risk and protective social factors with adolescent depression. However, this approach is not exhaustive, as factors such as ethnicity, religion, cultural influences, and specific neighborhood characteristics, in addition to bio-psychological factors, can also contribute to adolescent depression. Additionally, while this review provides a comprehensive examination of social factors and their association with adolescent depression, it does not establish causal explanations.

The review prioritizes the family domain over school and community factors, with the limited focus on neighborhood characteristics (only two studies) highlighting a gap in understanding their role in adolescent depression in LMICs. Furthermore, the predominant use of quantitative research methods limits the discussion of adolescents’ lived experiences and perspectives regarding their depressive experiences. The overrepresentation of studies from China may also introduce bias and limit the generalizability of findings to other LMICs.

Therefore, future research should employ diversified research methods, include studies from a wider range of LMICs to enhance generalizability, incorporate more research on the community domain, and utilize qualitative research to gain a deeper understanding of adolescents’ experiences with depression.

## Open access

This is an open-access article distributed under the terms of the Creative Commons Attribution License, which permits unrestricted use, distribution, and reproduction in any medium, provided the original author and source are credited.

## Author Contributions

IS: Conceptualization, methodology, data searching, screening and extraction, writing – original draft, and writing – review and editing.

SG: Data searching, screening and extraction, and writing – review and editing.

KE: Funding acquisition and writing – review and editing.

CK: Writing – review and editing.

PA: Funding acquisition and writing – review and editing.

All authors approved the final draft.

## Author disclaimer

The views expressed in this publication are those of the authors and not necessarily those of the NIHR or the UK government, the Court of the University of Aberdeen, the Board of Directors of the University of Rwanda, the Board of Directors of Addis Ababa University, the Board of Directors of The Sanctuary, or our International Advisory Board.

## Funding

This research was funded by the NIHR (NIHR133712) using UK international development funding from the UK Government to support global health research. The views expressed in this publication are those of the authors and not necessarily those of the NIHR or the UK government, the Court of the University of Aberdeen, the Board of Directors of the University of Rwanda, the Board of Directors of Addis Ababa University, the Board of Directors of The Sanctuary, or our International Advisory Board.

## Ethical Considerations

Ethical consent was not required

## Consent for Publication

Not applicable.

## Data Availability

All data produced in the present work are contained in the manuscript

## Notes

### Competing Interest Statement

The authors have declared no competing interest.

