## supplemental table for "The Social Determinants of Depression among Adolescents in Low- and Middle-Income Countries: A Scoping Review": Supplementary Materials for medrixiv.pdf

| Author, year | Title | Year | Country | Research Type | Research Approach | Sampling method | Study Characteristics | Number of participants | Data Collection Methods | Screening Tool | Findings |
|---|---|---|---|---|---|---|---|---|---|---|---|
| Alex et al., 2023 | A study on prevalence and risk factors of depression among adolescent girls studying in government and private schools-A comparative study | 2023 | India | Cross-sectional | Quantitative research | Probability sampling | Two private and two government run school adolescents | 391 | Questionnaire | Kutcher Adolescent Depression Scale (KADS) | Students from nuclear families exhibited a higher risk of depression. Among female students, those attending private schools were more likely to experience depression compared to their peers in government schools. In private schools, depression was primarily associated with academic stress, peer pressure, conflicts with friends, excessive smartphone use, and family discord. In contrast, in government schools, depression was mainly linked to family discord and the chronic illness of family members. |
| Shi et al., 2023 | A study on the correlation between family dynamic factors and depression in adolescents | 2023 | China | Cross-sectional | Quantitative research | Non-probability sampling | Primary school going adolescents | 4109 | Questionnaire | Self-Rating Depression Scale (SDS) | Improved family dynamics, marked by a positive family atmosphere and individuation, were associated with lower levels of depression. |
| Jayanthi et al., 2015 | Academic Stress and Depression among Adolescents: A Cross-sectional Study | 2015 | India | Cross-sectional | Quantitative research | Probability sampling | Adolescents at higher secondary schools | 1120 | Questionnaire | Beck Depression Inventory (BDI) | Adolescents experiencing academic stress were 2.4 times more likely to develop depression compared to those not experiencing academic stress. |
| Shabab et al., 2022 | Adolescent perspectives on depression as a disease of loneliness: a qualitative study with youth and other stakeholders in urban Nepal | 2022 | Nepal | Cross-sectional | Qualitative research | Non-probability sampling | adolescent with a history of depression and with no history of mental health conditions | 72 | Interview; Focus group discussion | Not mentioned | Loneliness is a key component of adolescent depression, emphasizing the need for culturally sensitive diagnostic criteria and interventions targeting loneliness. Adolescents distinguish depression from stress associated with family issues, peer relationships, and social media influences. |
| Pan et al., 2022 | After-School Extracurricular Activities Participation and Depressive Symptoms in Chinese Early Adolescents: Moderating Effect of Gender and Family Economic Status | 2022 | China | Panel/longitudinal | Quantitative research | Probability sampling | Junior high school students | 9672 | Questionnaire | PHQ-9 and CES-D | Time spent on homework, extracurricular tutoring, and playing online games was positively associated with depressive symptoms, while physical exercise was negatively associated. Gender and family economic status moderated these effects, with physical exercise showing a protective role against depression. |
| Liu et al., 2023 | Association between Child Abuse, Depression, and School Bullying among Chinese Secondary School Students | 2023 | China | Cross-sectional | Quantitative research | Probability sampling | Secondary school students | 3059 | Questionnaire | Zung SDS | Male students were more represented in the bully-victim group and experienced higher levels of physical and mental abuse during childhood. Female students were less likely to be involved in school bullying, but childhood abuse significantly increased their risk of depression. |
| Naveed et al., 2019 | Association of bullying experiences with depressive symptoms and psychosocial functioning among school going children and adolescents | 2019 | Pakistan | Cross-sectional | Quantitative research | Probability sampling | School-going children | 452 | Questionnaire | PHQ | It found that 28.8% of individuals reported victimization, and 32.3% reported perpetrating bullying. Both victims and bullies experienced negative emotional and social impacts, with those involved in both bullying and victimization showing the most severe depressive symptoms. Victimization was the strongest predictor of depressive symptoms, followed by perpetration and victimization. Age was linked only to bully perpetration. |
| Ferrajae et al., 2022 | Defense mechanisms mediate associations between exposure to adverse childhood experiences and anxiety and depression in Kenyan adolescents | 2022 | Kenya | Cross-sectional | Quantitative research | Probability sampling | Boarding school children | 475 | Questionnaire | The Trauma Symptom Checklist | Higher direct exposure to adverse childhood experiences (ACEs) was associated with increased depressive symptoms. Female adolescents reported greater exposure to both direct and indirect ACEs compared to males. Immature defense mechanisms mediated these associations. |
| Kaur et al., 2014 | Depression Among Adolescents in Relation to Their Academic Stress | 2014 | India | Cross-sectional | Quantitative research | Probability sampling | Senior Secondary School Adolescents | 200 | Questionnaire | BDI-II | A significant positive correlation was observed between academic stress and depression. Although girls experienced higher levels of academic frustration than boys, no significant gender differences were found in depression levels. |
| Bach and Louw et al., 2010 | Depression and exposure to violence among Venda and Northern Sotho adolescents in South Africa | 2010 | South Africa | Cross-sectional | Quantitative research | Probability sampling | High school students | 337 | Questionnaire | CDI | Girls had a higher prevalence of depression, while Venda adolescents experienced higher victimization rates than their Northern Sotho counterparts. Depression was more common among Northern Sotho boys than Venda boys, and exposure to violence was strongly correlated with depression across all groups. |
| Ma et al., 2022 | Depressive symptoms prevalence, associated family factors, and gender differences: A national cohort study of middle school students in China | 2020 | China | Panel/longitudinal | Quantitative research | Probability sampling | school going adolescents | 9869 | Questionnaire | 4-item CEPS-constructed scale | Depressive symptoms were found in 17.9% of 7th graders and 25.7% of 9th graders, with higher rates in girls. Positive parent-child relationships, particularly mother-child and father-child closeness, were associated with reduced depressive symptoms, especially in girls. The gender difference in depression prevalence was linked to significant increases in feelings of unhappiness and meaninglessness among girls. |
| Hua and Liu et al., 2021 | Dramaturgical perspective mediates the association between parenting by lying in childhood and adolescent depression and the protective role of parent-child attachment | 2021 | China | Cross-sectional | Quantitative research | Probability sampling | Secondary school adolescents | 964 | Questionnaire | Depression Anxiety Stress Scale (DASS) | Parenting by lying was positively associated with adolescent depression, with this relationship mediated by the adolescent's tendency to adopt a dramaturgical perspective—adjusting self-presentation across different social contexts. However, strong parent-child attachment functioned as a protective factor, mitigating the association between parenting by lying and adolescent depression. |
| Zhang et al., 2022 | Effects of Father-Adolescent and Mother-Adolescent Relationships on Depressive | 2022 | China | Panel/longitudinal | Quantitative research | Probability sampling | Middle school adolescents | 8794 | Questionnaire | Four items to measure adolescents | The study found that stronger parent-child relationships are associated with fewer depressive symptoms, particularly for boys, where close father-adolescent relationships correlate with |

| Author, year | Title | Year | Country | Research Type | Research Approach | Sampling method | Study Characteristics | Number of participants | Data Collection Methods | Screening Tool | Findings |
|---|---|---|---|---|---|---|---|---|---|---|---|
| | Symptoms among Chinese Early Adolescents | | | | | | | | | depressive symptoms | lower depression levels. Additionally, adolescents from wealthier families tend to have better parent-child relationships and are less likely to experience depression. |
| Xia et al., 2022 | Effects of negative life events on depression in middle school students: The chain-mediating roles of rumination and perceived social support | 2022 | China | Cross-sectional | Quantitative research | Non-probability sampling | Middle school adolescents | 493 | Questionnaire | CDI | Depression showed a stronger association in females than males, with negative life events directly influencing depressive symptoms and indirectly contributing to increased rumination and decreased perceived social support, further elevating depression levels. |
| Guo et al., 2014 | Effort-Reward Imbalance at School and Depressive Symptoms in Chinese Adolescents: The Role of Family Socioeconomic Status | 2014 | China | Cross-sectional | Quantitative research | Probability sampling | Middle and high school adolescents | 1774 | Questionnaire | CES-DC | Low family SES and high school-related stress were associated with depressive symptoms among adolescents. The prevalence of school-related stress and depressive symptoms was significantly higher in adolescents from low SES families compared to those from high SES families. Students from low SES families were also significantly older and smoked more frequently. |
| Yi et al., 2013 | Exposure to violence in relation to depressive symptoms among male and female adolescent students in Cambodia | 2013 | Cambodia | Cross-sectional | Quantitative research | Probability sampling | High school adolescents | 1943 | Questionnaire | The Asian Adolescent Depression Scale | Depressive symptoms were linked to victimization or witnessing violence, with witnessing community violence significantly affecting only female students. Factors such as lower family income, poor school performance, and lack of family support were associated with higher depressive symptoms, especially in females. The findings highlight gender differences in the effects of violence exposure on adolescent mental health. |
| Huang et al., 2022 | Family functioning and adolescent depression: A moderated mediation model of self-esteem and peer relationships | 2022 | China | Cross-sectional | Quantitative research | Probability sampling | Middle school adolescents | 562 | Questionnaire | The youth self-rating index (YSR)-the depression subscale | Family functioning negatively predicts adolescent depression, with self-esteem mediating this relationship. Peer relationships were found to moderate the effect of self-esteem on depression. |
| Xu et al., 2019 | Family Socioeconomic Status and Adolescent Depressive Symptoms in a Chinese Low- and Middle-Income Sample: The Indirect Effects of Maternal Care and Adolescent Sense of Coherence | 2019 | China | Cross-sectional | Quantitative research | Probability sampling | Public middle and high school adolescents | 783 | Questionnaire | CES-DC | SES, maternal care, and adolescent sense of coherence were positively interrelated and negatively associated with adolescent depressive symptoms. SES influenced adolescent depressive symptoms indirectly through maternal care alone and sequentially through both maternal care and sense of coherence.. |
| Peng et al., 2021 | Father-Child Conflict and Chinese Adolescent Depression: A Moderated Mediation Model | 2021 | China | Cross-sectional | Quantitative research | Probability sampling | Middle-school students | 654 | Questionnaire | CES-DC | Father-child conflict was positively associated with adolescent depression and negatively correlated with RESE. Both positive and negative emotional regulation self-efficacy partially mediated the relationship between father-child conflict and depression. Gender moderated this relationship, with girls being more affected by low levels of negative emotional regulation self-efficacy. Additionally, father-child conflict was significantly lower in boys than in girls. |
| Zou et al., 2020 | Higher Socioeconomic Status Predicts Less Risk of Depression in Adolescence: Serial Mediating Roles of Social Support and Optimism | 2020 | China | Cross-sectional | Quantitative research | Probability sampling | Junior and senior high school adolescents | 652 | Questionnaire | CES-DC | Socioeconomic status (SES) negatively predicted adolescent depression, with social support and optimism mediating this relationship. Higher SES was associated with greater social support and increased optimism, which in turn reduced depressive symptoms. |
| Wang et al., 2024 | Interparental Conflict and Early Adolescent Depressive Symptoms: Parent-Child Triangulation as the Mediator and Grandparent Support as the Moderator | 2024 | China | Cross-sectional | Quantitative research | Probability sampling | Elementary and junior high school adolescents | 761 | Questionnaire | CES-DC | Adolescents are at greater risk of depression when exposed to frequent parental conflict, particularly if they are caught in the middle due to unstable or stable coercive coalitions. Girls and those in higher grades are more vulnerable. However, support from grandparents can mitigate the negative effects of both interparental conflict and unstable coercive coalitions on the child's depression. |
| He et al., 2019 | Interpersonal Conflict, School Connectedness and Depressive Symptoms in Chinese Adolescents: Moderation Effect of Gender and Grade Level | 2019 | China | Cross-sectional | Quantitative research | Probability sampling | Primary and secondary school adolescents | 6576 | Questionnaire | Depression Self-Rating Scale for Children (DSRSC) | Conflicts with parents, teachers, and peers were associated with increased depressive symptoms in adolescents, while school connectedness had a protective effect. Gender and grade level moderated these associations, with stronger effects observed in females and primary school students. |
| Emery et al., 2022 | Invasive Exploitation and the Multiplicative Hypothesis: Polyvictimization and Adolescent Depression Symptoms in Nepal | 2022 | Nepal | Cross-sectional | Quantitative research | Probability sampling | Mother-adolescent dyads | 565 | Questionnaire | CES-D | Reports of physical abuse, neglect, and sexual abuse remain high. Both neglect and sexual abuse were positively associated with adolescent depressive symptoms, with older adolescents exhibiting more symptoms. |
| Gao et al., 2022 | Life events and depression among children and adolescents in southwest China: a two-stage moderated mediation model of social support and cognitive styles | 2022 | China | Cross-sectional | Quantitative research | Probability sampling | Primary and secondary school adolescents | 2804 | Questionnaire | CDS | Negative life events were linked to increased depression, with social support partially mediating this relationship. Cognitive styles moderated these effects, as positive styles buffered the impact of life events and strengthened social support's protective role, while negative styles intensified life events' effects and weakened social support's benefits. |
| Vasconcelos et al., 2020 | Life satisfaction mediates the association between childhood maltreatment and depressive symptoms: a study in a sample of Brazilian adolescents | 2020 | Brazil | Cross-sectional | Quantitative research | Probability sampling | Adolescents in public schools | 342 | Questionnaire | CDI | Childhood maltreatment was negatively correlated with life satisfaction and positively associated with depressive symptoms. Life satisfaction partially mediated this relationship, indicating that lower life satisfaction may exacerbate the impact of maltreatment on depressive symptoms. |
| Liu et al., | Mediating effect of social support on the | 2020 | China | Cross- | Quantitative | Probability | Secondary school | 1512 | Questionnaire | CDI | Negative life events, when coupled with low social support, contribute to an increased risk of |

| Author, year | Title | Year | Country | Research Type | Research Approach | Sampling method | Study Characteristics | Number of participants | Data Collection Methods | Screening Tool | Findings |
|---|---|---|---|---|---|---|---|---|---|---|---|
| 2020 | association between life events and depression: A cross-sectional study of adolescents in Chongqing China | | | sectional | research | sampling | adolescents | | | | depression. The frequency and perceived importance of social support mediate the relationship between life events and depression. Additionally, ethnicity is significantly associated with depressive symptoms. |
| Song et al., 2022 | Mediating effects of parent-child relationship on the association between childhood maltreatment and depressive symptoms among adolescents | 2022 | China | Cross-sectional | Quantitative research | Probability sampling | Middle school students | 14500 | Questionnaire | CDI | Childhood maltreatment weakens the parent-child relationship, contributing to increased depressive symptoms. This effect is more pronounced in girls and adolescents with siblings compared to boys and only children. |
| Jayanthi and Thirunavukarasu et al., 2016 | Perceived Social Support - A Risk Factor for Depression Among Adolescents: An Analytical Study | 2016 | India | Cross-sectional | Quantitative research- A community based case control study | Probability sampling | High school students | 1120 | Questionnaire | BDI | Adolescents with lower levels of perceived social support were 1.9 times more likely to develop depression compared to those with adequate support. Both case and control groups reported stronger family support than support from peers or significant others, though the case group's family support was weaker. Notably, adolescents with depression had higher perceived social support overall. These findings highlight the critical role of support from parents and peers in predicting future depression in adolescents. |
| Miloseva et al., 2017 | Perceived social support as a moderator between negative life events and depression in adolescence: implications for prediction and targeted prevention | 2017 | Macedonia | Cross-sectional-Corelational study | Quantitative research | Probability sampling | Adolescents from psychiatric clinics, primary and secondary schools | 412 | Questionnaire | CES-D | The link between depressive symptoms and negative life events depended on the level of perceived social support. The moderating effect was significant only in the subclinical group, indicating that social support may be especially important for adolescents with subclinical depressive symptoms. These findings suggest that enhancing social support could serve as an effective preventive strategy against depression in adolescents. |
| Chekol et al., 2023 | Predictors of depression among school adolescents in Northwest, Ethiopia, 2022: institutional based cross-sectional | 2023 | Ethiopia | Cross-sectional | Quantitative research | Probability sampling | Public and private high school adolescent students | 584 | Questionnaire | PHQ-9 | Female gender, small family size, history of alcohol use, attendance at public schools, and a history of abuse were associated with a higher likelihood of experiencing depression. |
| Singh et al., 2023 | Prevalence and Determinants of Depressive Symptoms among Young Adolescents in Malaysia: A Cross-Sectional Study | 2023 | Malaysia | Cross-sectional | Quantitative research | Probability sampling | School going adolescents | 1350 | Questionnaire | CESD | Female gender, smoking, being bullied, feelings of loneliness, and lack of parental supervision were significant determinants of depressive symptoms. |
| Ibrahim et al., 2017 | Prevalence and predictors of depression and suicidal ideation among adolescents attending government secondary schools in Malaysia | 2017 | Malaysia | Cross-sectional | Quantitative research | Probability sampling | High school students | 1800 | | PHQ-9 | Adolescents involved in bullying—both victims and perpetrators—and those who smoke exhibited higher rates of depression. Depression was more prevalent among females and significantly associated with bullying, smoking, anxiety, and stress. Additionally, suicidal ideation was markedly more common among adolescents with depression than those without. |
| Sun et al., 2021 | Psycho-social factors associated with high depressive symptomatology in female adolescents and gender difference in adolescent depression: an epidemiological survey in China's Hubei Province | 2021 | China | Cross-sectional | Quantitative research | Probability sampling | School going adolescents | 4100 | Questionnaire | Chinese version of CESD | High levels of depressive symptoms were more prevalent among females and significantly associated with older age, single-parent families, and low paternal education. Adolescents with fathers who had only primary education or less, and those from single-parent households, showed higher odds of depression. Conversely, regular physical activity was linked to lower odds of depressive symptoms. |
| Shen et al., 2022 | Reciprocal associations between peer victimization and depressive symptoms among Chinese children and adolescents: Between and within-person effects | 2022 | China | Panel/longitudinal | Quantitative research | Probability sampling | primary school going adolescents | 1205 | Questionnaire | CES-DS | Peer victimization predicts increased depressive symptoms, while experiencing depressive symptoms increases the likelihood of subsequent victimization, regardless of gender, age, or socioeconomic background. |
| Yee & Sulaiman 2017 | Resilience as Mediator in the Relationship between Family Functioning and Depression among Adolescents from Single Parent Families | 2017 | Malysia | Cross-sectional | Quantitative research | Purposive sampling | Secondary school adolescents | 232 | Questionnaire | BDI-II | Resilience mediated the relationship between family adaptability and depression and partially mediated that between family cohesion and depression. Strong family bonds provide crucial social support, fostering resilience in adolescents. Thus, family cohesion more strongly relates to resilience than family adaptability. |
| Wang et al., 2023 | Risk and Protective Factors of Depression in Family and School Domains for Chinese Early Adolescents: An Association Rule Mining Approach | 2023 | China | Cross-sectional | Quantitative research | Probability sampling | Primary and secondary school adolescents | 2455 | Questionnaire | The Depression Self-Rating Scale for Children | Family cohesion and conflict were key factors in adolescent depression, with adverse effects heightened among females, middle school students, and those from disadvantaged backgrounds. Limited school mental health resources further exacerbated the risk. |
| Mohanraj et al., 2010 | Risk and Protective Factors to Depressive Symptoms in School-Going Adolescents | 2010 | India | Cross-sectional | Quantitative research | Probability sampling | School going adolescents | 964 | Questionnaire | (BDI | Self-esteem is the strongest protective factor against depression, followed by peer acceptance, family relationships, and social support. In contrast, daily hassles were found to be a stronger risk factor for depression than life events. These results emphasize the importance of positive personal, social, and family dynamics in reducing the risk of depression among adolescents. |
| Dervishi et al., 2019 | School bullying and symptoms of depression | 2019 | Albania | Cross-sectional | Quantitative research | Probability sampling | Primary school students | 284 | Questionnaire | (CDI | Bullying victimization is significantly positively correlated with depressive symptoms, as adolescents who frequently experience bullying are more likely to report depression, whereas prosocial behaviors are associated with fewer depressive symptoms. |
| Liu et al., 2022 | Social Support and Depressive Symptoms Among Adolescents During the COVID-19 Pandemic: The Mediating Roles of Loneliness and Meaning in Life | 2022 | China | Cross-sectional | Quantitative research | Probability sampling | High school students | 1317 | Questionnaire | BDI-II | Social support had both direct and indirect effects on depressive symptoms. Directly, social support reduced depressive symptoms, while indirectly, it influenced depressive symptoms by reducing loneliness and enhancing the sense of meaning in life. |
| Dhamaya | The association of depression with child abuse | 2020 | Indonesia | Cross- | Quantitative | Probability | Junior high | 786 | Questionnaire | CDI | The study found that psychological victimization, exposure to violence, physical victimization, |

| Author, year | Title | Year | Country | Research Type | Research Approach | Sampling method | Study Characteristics | Number of participants | Data Collection Methods | Screening Tool | Findings |
|---|---|---|---|---|---|---|---|---|---|---|---|
| nti et al., 2020 | among Indonesian adolescents | | | sectional | research | sampling | school students | | | | neglect, and sexual victimization were all associated with an increased risk of depression, with psychological violence posing the highest risk, followed by exposure to and physical violence, while sexual violence was less prevalent. |
| Chung et al., 2020 | The association of perceived neighbourhood factors and social class with depressive symptoms among Grade 6 elementary school children in Jamaica | 2020 | Jamaica | Cross-sectional | Quantitative research | Probability sampling | Grade six public and private school adolescents | 321 | Questionnaire | The Adolescent Depression Rating Scale (ADRS) | Neighborhood factors, such as social networks and attachment, were negatively associated with depression, while poor quality and crime showed a positive association. These factors may mediate the link between low socioeconomic status and depression, with lower status, weaker networks, and poorer quality correlating with higher depression scores. |
| Qin et al., 2021 | The Impact of Academic Achievement and Parental Practices on Depressive Symptom Trajectories Among Chinese Adolescents | 2021 | China | Cross-sectional | Quantitative research | Probability sampling | Public general middle schools adolescents | 2576 | Questionnaire | CDI | Adolescents who perform well academically and receive parental support for independence have a lower risk of developing depressive symptoms, whereas those experiencing parental academic pressure are more likely to exhibit higher depressive symptoms. |
| Khasakhala et al., 2012 | The prevalence of depressive symptoms among adolescents in Nairobi public secondary schools: association with perceived maladaptive parental behaviour | 2012 | Kenya | Cross-sectional | Quantitative research | Probability sampling | School going adolescents | 1276 | Questionnaire | CDI | Depressive symptoms increased with age (14–17 years) and were more prevalent among adolescents who perceived maternal rejection, lacked emotional attachment from parents, or experienced under protective paternal behavior. Female adolescents exhibited higher depressive symptoms and suicidal behavior, with boarding students experiencing more symptoms than day students. |
| Wang et al., 2022 | The Relationship Between Parent-Adolescent Communication and Depressive Symptoms: The Roles of School Life Experience, Learning Difficulties and Confidence in the Future | 2022 | China | Panel/longitudinal | Quantitative research | Probability sampling | Primary school going students | 9449 | Questionnaire | Feeling of four item scales for the past seven days | Positive father-adolescent communication, favorable school experiences, and greater confidence in the future were identified as protective factors against depressive symptoms in adolescents. In contrast, depressive symptoms were associated with perceived maternal rejection, weak emotional attachment to both parents, under-protective parenting, and learning difficulties. |
| Tsehay et al., 2020 | The Role of Adverse Childhood Experience on Depression Symptom, Prevalence, and Severity among School Going Adolescents | 2020 | Ethiopia | Cross-sectional | Quantitative research | Probability sampling | Secondary school adolescents | 546 | Questionnaire | PHQ-9 | Higher ACE exposure increasing the risk of depression. Male gender, stronger social support, and lower grade levels were associated with reduced depressive symptoms. |
| Zhang et al., 2022 | The role of family and peer factors in the development of early adolescent depressive symptoms: A latent class growth analysis | 2022 | China | Panel/longitudinal | Quantitative research | Probability sampling | Primary and middle school adolescents | 586 | Questionnaire | CES-D | Female adolescents and those experiencing psychological aggression, neglect, high parental psychological control, victimization through traditional bullying and cyber bullying, and poor-quality friendships were more likely to belong to the increasing depressive symptoms class. |
| Li et al., 2022 | The role of parental conflict in predicting adolescent depression symptoms during the COVID-19 pandemic: A longitudinal study | 2022 | China | Panel/longitudinal | Quantitative research | Probability sampling | Secondary school adolescents | 655 | Questionnaire | PHQ-9 | It found a double-hump pattern in depression, peaking at W1 and W3, with parental conflict, especially in content and resolution, being a significant factor. Parental conflict increased depression both directly and indirectly by reducing family support and enhancing feelings of burdensomeness. The study emphasizes the importance of addressing family dynamics, particularly parental conflict, as a key risk factor in adolescent depression during challenging times. |
| Lakhdir et al., 2021 | The role of parent-to-child maltreatment in the pathway of self-reported depressive symptoms in Pakistani adolescents | 2021 | Pakistan | Panel/longitudinal | Quantitative research | Probability sampling | Adolescents in the community | 751 | Questionnaire | CES-D | Maltreated adolescents reported depressive symptoms, with females showing higher rates. Children with uneducated parents, stressful home environments, and those without formal education were at a higher risk. |

Supplementary materials III- Preferred Reporting Items for Systematic reviews and Meta-Analyses extension for Scoping Reviews (PRISMA-ScR) Checklist

| SECTION | ITEM | PRISMA-ScR CHECKLIST ITEM | REPORTED ON PAGE # |
|---|---|---|---|
| **TITLE** | | | |
| Title | 1 | The Social Determinants of Depression among Adolescents in Low- and Middle-Income Countries: A Scoping Review | 1 |
| **ABSTRACT** | | | |
| Structured summary | 2 | This scoping review, guided by Arksey and O'Malley's framework, synthesizes findings from 48 studies to examine the social determinants of adolescent depression in low- and middle-income countries (LMICs), with particular emphasis on family, school, and neighborhood factors. The review highlights several key risk factors for adolescent depression, including age, female gender, family conflict, harsh parenting, lower socioeconomic status, maltreatment, academic stress, lack of peer acceptance, bullying, excessive tutoring, and neighborhood violence, while also recognizing social support as a significant protective factor. However, the predominant shares of the studies were cross-sectional and concentrated in Asia, especially China, leaving underrepresented regions such as Africa, Europe, and South America. Research on neighborhood-level factors was sparse, and the experiences of out-of-school adolescents were neglected. Moreover, not only was the interplay between family, school, and neighborhood influences on adolescent depression insufficiently explored, but limitations were also observed in exploring each domain more broadly. Therefore, addressing these gaps is critical for the development of more targeted and effective interventions that can address the unique challenges faced by adolescents in various cultural and socioeconomic contexts. | 1 |
| **INTRODUCTION** | | | |
| Rationale | 3 | Adolescent depression is increasingly recognized as a significant public health concern in low- and middle-income countries (LMICs). This transitional stage of life is marked by profound biological, psychological, and social changes that heighten vulnerability to mental health challenges, including depression. While much of the existing research has centered on psychological and biological factors, the social determinants influencing adolescent depression in LMICs remain insufficiently explored. Understanding how family, school, and neighborhood environments shape adolescent depression is crucial for gaining a holistic perspective on its social determinants. However, a broader and more detailed analysis of each domain, as well as the interplay between these factors, is notably lacking. This scoping review aims to address this gap by systematically mapping the available evidence on the social determinants of adolescent depression and identifying evidence gaps that require further investigation. | 2-4 |
| Objectives | 4 | To examine the social determinants of adolescent depression in LMICs, focusing on risk and protective factors within family, school, and neighborhood domains while identifying gaps in the existing research | 4 |
| **METHODS** | | | |
| Protocol and registration | 5 | The protocol for this scoping review has been pre-published on medRxiv with the registration number **2024.10.04.24314891v1** and can be accessed at | 4 |

| SECTION | ITEM | PRISMA-ScR CHECKLIST ITEM | REPORTED ON PAGE # |
|---|---|---|---|
| | | https://www.medrxiv.org/content/10.1101/2024.10.04.24314891v1.full. | |
| Eligibility criteria | 6 | This review included empirical studies published in English from January 2010 to December 2023, focusing on depression among adolescents in LMICs. Studies employing any methodology were eligible if they involved a general adolescent population (excluding those with diagnosed biological or psychological conditions) and examined at least one social factor influencing depression. | 5 |
| Information sources* | 7 | The primary databases searched included Google Scholar, PsychINFO (via Ovid), Web of Science, PubMed, Sociological Abstracts (via ProQuest), and Embase (via Ovid). | 5 |
| Search | 8 | We conducted the literature search using advanced search functionalities within the target databases. We employed pre-specified search terms (outlined in the protocol) to retrieve relevant articles, which we then exported into EndNote for management and analysis. | 5 |
| Selection of sources of evidence† | 9 | An initial search using terms approved by PA, IW, and SG retrieved 48,468 records. After removing 42,045 duplicates with Covidence, 6,417 records remained for title and abstract screening. IW and SG excluded 5,964 irrelevant records, leaving 453 for full-text review. Additionally, 16 studies were identified through manual searches and directly included in the full-text assessment. Before the review, IW and SG independently screened a sample of records to ensure consistent eligibility application, achieving a 94% inter-rater agreement. Discrepancies were resolved through discussion. Of the 453 potentially relevant articles, 48 were included. These were uploaded into NVivo Version 14, and IW and SG independently coded a 10% sample. After feedback from PA and KE, they coded the remaining records. Regular discussions addressed coding discrepancies and ensured consistency and alignment in the coding approach. | 4-5 |
| Data charting process‡ | 10 | Key data, including study design, sampling method, data collection, and findings, were extracted by IW and verified by SG, KE, and CK. | 5 |
| Data items | 11 | Data extracted included: Age, Gender, Other characteristics, Family size and structure, Parental characteristics, Family dynamics, Life events and maltreatment, Parenting style, Socio-economic status, Academic stress, School bullying, Relation with peers, Relation with teachers, School types, Violence and crime, and findings related to depression. | 8-15 |
| Critical appraisal of individual sources of evidence§ | 12 | No formal quality assessment was conducted; however, the included studies were published in peer-reviewed journals. | 5 |
| **RESULTS** | | | |
| Selection of sources of evidence | 14 | The sources of evidence for this scoping review were selected based on their relevance to adolescent depression and their inclusion of factors related to individual, family, and school environments. Studies were chosen through systematic database searches, with an emphasis on peer-reviewed articles, recent publications, and research conducted in low- and middle-income countries. The selection criteria ensured that only studies with a clear focus on adolescent depression and its correlates were included. | 5-6 |
| Characteristics of sources of | 15 | The included studies varied in their designs, including cross-sectional, longitudinal, and qualitative research. Most | 7-8 |

| SECTION | ITEM | PRISMA-ScR CHECKLIST ITEM | REPORTED ON PAGE # |
|---|---|---|---|
| evidence | | studies were conducted in school and community settings, focusing on adolescents aged 12-18. They explored a range of individual characteristics (e.g., age, gender), family-related factors (e.g., family structure, parental education, family conflict), and school-related factors (e.g., academic pressure, bullying, peer relationships). | |
| Critical appraisal within sources of evidence | 16 | No formal quality assessment was conducted; however, the included studies were published in peer-reviewed journals. | 5 |
| Results of individual sources of evidence | 17 | The findings from individual sources consistently identified age and gender as significant predictors of depressive symptoms, with older adolescents and girls showing higher levels of depression. Family-related factors, including family type, parental education, family conflict, and abuse, were strongly linked to depressive symptoms, particularly among girls. Positive family relationships were found to be protective, while negative family dynamics, such as parental conflict or maltreatment, were risk factors. In school environments, academic pressure, bullying, and peer relationships were significant contributors to depression, with bullying victimization and lack of peer acceptance increasing the risk. Positive teacher relationships and peer support were identified as protective factors that mitigated depressive symptoms. These results underscore the importance of a multifaceted approach to addressing adolescent depression, taking into account personal, familial, and school-related influences. | 8-15 |
| Synthesis of results | 18 | The studies reviewed highlight the multifaceted nature of adolescent depression, with individual, family, and school factors playing key roles. Age and gender were significant predictors, with older adolescents and girls experiencing higher levels of depression. Family dynamics, including family type, parental education, conflict, and abuse, were strongly associated with depression, particularly in girls. Supportive family relationships acted as protective factors, while negative dynamics increased risk. School-related factors such as academic pressure, bullying, and peer relationships were also significant contributors to depressive symptoms. Positive teacher relationships and peer support served as protective factors. This review emphasizes the need for interventions targeting individual, familial, and school domains, particularly in fostering family support, healthy peer relationships, and balanced academic expectations. | 8-15 |
| **DISCUSSION** | | | |
| Summary and conclusion of evidence | 19 | This review examined the association between social factors and adolescent depression, focusing on low- and middle-income countries (LMICs). It synthesized findings from 51 studies, revealing that family-related factors were the most studied (76%), followed by school-related factors (48.1%), while neighborhood-related factors were minimally explored. The review identified significant regional and methodological gaps, with most studies conducted in Asia, particularly China, and limited representation from Africa, Europe, and South America. Most studies were cross-sectional, and only one qualitative study was included, highlighting a need for more diverse research methodologies. Furthermore, the focus on school-going adolescents excluded the experiences of out-of-school youth. The literatures have narrow focus on external social stressors without addressing underlying structural factors, such as socioeconomic social structural factors, which are crucial in shaping these stressors and contributing to adolescent depression. It | 16-19 |

| SECTION | ITEM | PRISMA-ScR CHECKLIST ITEM | REPORTED ON PAGE # |
|---|---|---|---|
| | | emphasizes the importance of examining the interplay of multiple social factors and stressors to develop a more comprehensive understanding of adolescent depression in LMICs. | |
| Limitations | 20 | The review focused on risk and protective social factors in adolescent depression but acknowledged that it does not provide an exhaustive view, as factors such as ethnicity, religion, cultural influences, specific neighborhood characteristics, and bio-psychological elements may also play a role. While the review offers a comprehensive examination of social factors, it does not establish causal relationships. The study prioritizes family-related factors over school and community factors, with a particularly limited focus on neighborhood characteristics, which underscores a gap in understanding their role in adolescent depression in LMICs. Additionally, the reliance on quantitative methods restricts the exploration of adolescents' lived experiences with depression. The overrepresentation of studies from China introduces potential bias, limiting the generalizability of findings to other LMICs. The review calls for future research to employ diverse methodologies, include a broader range of LMICs for better generalizability, explore the community domain more extensively, and incorporate qualitative research to gain a more nuanced understanding of adolescents' depressive experiences. | 19 |
| **FUNDING** | | | |
| Funding | 22 | This research was funded by the NIHR (NIHR133712) using UK international development funding from the UK Government to support global health research. The views expressed in this publication are those of the authors and not necessarily those of the NIHR or the UK government, the Court of the University of Aberdeen, the Board of Directors of the University of Rwanda, the Board of Directors of Addis Ababa University, the Board of Directors of The Sanctuary, or our International Advisory Board | 21 |

Supplementary Materials IV: Countries by Continent and Income

| Country | No. of studies | Continent | Income Level |
|---|---|---|---|
| Albania | 1 | Europe | Upper-middle-income |
| Brazil | 1 | South America | Upper-middle-income |
| Cambodia | 1 | Asia | Lower-middle-income |
| China | 25 | Asia | Upper-middle-income |
| Ethiopia | 2 | Africa | Low-income |
| India | 5 | Asia | Lower-middle-income |
| Indonesia | 1 | Asia | Lower-middle-income |
| Jamaica | 1 | North America | Upper-middle-income |
| Kenya | 2 | Africa | Lower-middle-income |
| Malaysia | 3 | Asia | Upper-middle-income |
| Nepal | 2 | Asia | Low-income |
| Pakistan | 2 | Asia | Lower-middle-income |
| South Africa | 1 | Africa | Upper-middle-income |
